# Identification of amino acid metabolism-related biomarkers in liver fibrosis: a transcriptomic analysis with experimental validation

**DOI:** 10.64898/2026.05.17.26353417

**Authors:** Z Liu, X. Liu

## Abstract

**Background:** Liver fibrosis (LF) represents a pivotal pathological phase in the advancement of chronic liver disorders toward cirrhosis. Amino acid metabolism reprogramming plays a pivotal role in its pathogenesis, yet the underlying molecular mechanisms remain incompletely understood.

**Methods:** Integrating three public datasets (GSE14323, GSE84044, and GSE136103) with amino acid metabolism-related gene sets, we performed consensus clustering, machine learning algorithms, functional enrichment analysis, immune microenvironment composition, regulatory network construction, and drug prediction.

**Results:** Fibrotic samples were classified into two amino acid metabolism-related subtypes with distinct immune landscapes and functional phenotypes. Through integrated analysis of differentially expressed genes (DEGs) common to both subtypes, fibrotic versus control comparisons, and amino acid metabolism–related gene sets, four biomarkers -- GSTP1, LDHB, OXCT1, and PTGDS--were identified. These biomarkers were enriched in pathways related to epithelial–mesenchymal transition, interferon responses, and TNFα/NF-κB signaling. Notably, GSTP1 and LDHB positively correlated with M1 macrophage infiltration and negatively with regulatory T cell abundance. Single-cell transcriptomic analysis revealed that cholangiocytes expressed all four biomarkers with elevated levels in fibrosis and interacted with macrophages/mesenchymal cells via MIF-CD74/CXCR4. Regulatory network analysis highlighted key modulators, including MALAT1, hsa-miR-3163, OXCT1, SMAD4, and RELA. Furthermore, 5-fluorouracil was predicted as a multi-target compound, with the strongest predicted binding affinity for OXCT1. *In vitro* validation confirmed the upregulation of GSTP1 and LDHB, aligning with the bioinformatics findings.

**Conclusion:** This study identified four amino acid metabolism-related biomarkers, revealing immune heterogeneity and cholangiocyte-centered intercellular communication in LF. These findings establish a foundation for biomarker-based diagnosis, subtype-guided patient stratification, and the development of cell-type-specific therapeutic strategies in LF.

## 1 Introduction

Liver fibrosis (LF) is a common pathological stage in the progressive course of various chronic liver diseases and serves as an important pathological basis for the development of cirrhosis, portal hypertension, liver failure, and even hepatocellular carcinoma (Akkız et al. 2024; Codotto et al. 2025; Devarbhavi et al. 2023; Parola and Pinzani 2024; Wiering et al. 2023; Younossi et al. 2023). Its etiology is complex and may be triggered by multiple factors, including viral hepatitis, alcohol-related liver disease, metabolic dysfunction-associated steatotic liver disease, and autoimmune liver disease (Akkız et al. 2024; Codotto et al. 2025; Devarbhavi et al. 2023; Parola and Pinzani 2024; Younossi et al. 2023). Regardless of the etiology, the core pathological mechanism of LF is an abnormal wound healing response caused by persistent liver injury and tissue repair disorders, which is characterized by activation of hepatic stellate cells, excessive deposition of extracellular matrix, and continuous remodeling of the liver microenvironment (Akkız et al. 2024; Codotto et al. 2025; Parola and Pinzani 2024; Wiering et al. 2023). During this process, injured hepatocytes, hepatic stellate cells, macrophages, cholangiocytes, endothelial cells, and diverse immune cell populations interact with each other to drive inflammatory amplification and fibrotic deposition; among them, classical pro-fibrotic signaling pathways such as TGF-β/Smad play pivotal roles in disease progression (Akkız et al. 2024; Codotto et al. 2025; Parola and Pinzani 2024; Wiering et al. 2023; Zhang et al. 2023). Although early-stage LF is reversible to some extent, current understanding of its early diagnosis, molecular classification, and precise therapeutic targets remains limited. Therefore, a deeper understanding of the key molecular basis and regulatory networks underlying LF is of great significance for improving precision diagnosis and individualized intervention strategies (Codotto et al. 2025; Parola and Pinzani 2024; Zhang et al. 2023).

In recent years, metabolic reprogramming has been increasingly recognized as an important mechanism that promotes tissue injury, inflammatory responses, and fibrotic progression, among which abnormalities in amino acid metabolism have attracted particular attention (Horn and Tacke 2024; Paulusma et al. 2022; Yang et al. 2023). Amino acids are not only essential for protein synthesis but also play critical roles in maintaining redox homeostasis, energy metabolism, immune regulation, and intracellular signaling (Paulusma et al. 2022; Yang et al. 2023). Emerging evidence has shown that dysregulated amino acid metabolism participates in the development of multiple liver diseases by influencing immune cell effector functions, inflammatory intensity, and fibrosis-related signaling pathways (Horn and Tacke 2024; Paulusma et al. 2022; Yang et al. 2023). For example, glutamine/ammonia metabolic reprogramming has been shown to be closely associated with the progression of LF, while branched-chain amino acid intervention has been reported to alleviate experimental LF by modulating the TGF-β1/Smad signaling pathway (Horn and Tacke 2024; Huang et al. 2025; Khedr and Khedr 2017). In addition, specific amino acid metabolism-related modifications may also contribute to hepatic fibrogenesis by regulating the inflammatory microenvironment and cell adhesion (Schonfeld et al. 2023). These findings suggest that amino acid metabolism-related genes may not only participate in the pathological process of LF but may also serve as potential diagnostic biomarkers and therapeutic targets (Horn and Tacke 2024; Huang et al. 2025; Khedr and Khedr 2017; Schonfeld et al. 2023; Yang et al. 2023). Nevertheless, systematic integrative analyses of amino acid metabolism-related molecular abnormalities in LF remain limited, and their molecular heterogeneity, immune relevance, and potential cellular origins have not yet been fully elucidated (Horn and Tacke 2024; Yang et al. 2023).

With the rapid development of high-throughput sequencing technologies, transcriptomic profiling has emerged as a powerful approach for identifying key molecular networks and potential biomarkers in LF (Codotto et al. 2025). Therefore, in this context, we utilized transcriptomic datasets from the Gene Expression Omnibus (GEO) database, combined with amino acid metabolism-related gene sets, to identify amino acid metabolism-related biomarkers in LF. Biomarkers were screened by integrating cluster analysis, differential expression analysis, and machine learning methods, and were preliminarily validated in in vitro cellular models. Subsequently, through combining immune infiltration analysis, regulatory network construction, and single-cell analysis, we systematically explored the potential mechanisms of action of these biomarkers in LF. This integrated strategy provides new insights into the molecular and immunological mechanisms of liver fibrosis and lays a theoretical foundation for the development of novel diagnostic and therapeutic strategies.

## 2 Materials and Methods

### 2.1 Data collection

The bulk RNA-Seq datasets GSE14323 comprising 60 liver tissue specimens—41 from LF patients and 19 from healthy controls (platform: GPL571) and GSE84044 including 124 samples—81 diseased and 43 control (platform: GPL570), as well as scRNA-seq dataset GSE136103 consisting of 20 liver tissue samples—10 per group (platform: GPL20301), were retrieved from the GEO database (https://www.ncbi.nlm.nih.gov/geo/). Furthermore, a curated list of 417 amino acid metabolism-related genes (AAMRGs) was extracted from the Molecular Signatures Database (MSigDB) (https://www.gsea-msigdb.org/gsea/msigdb).

### 2.2 Consensus clustering

Utilizing LF samples (n = 41) from the GSE14323 cohort and 417 AAMRGs, consensus clustering was executed with the ConsensusClusterPlus (v1.62.0) (Wilkerson and Hayes 2010). The partitioning around medoids (PAM) algorithm, coupled with Euclidean distance as the similarity metric, identified two distinct molecular subtypes among the samples.

### 2.3 Differential analysis and functional annotation

Differential gene expression analysis was implemented on the GSE14323 cohort via limma (v3.54.2) (Ritchie et al. 2015). Two distinct comparisons were carried out: (1) amino acid metabolism–related subgroups—specifically cluster2 versus cluster1 (|logLJ fold change (FC)| > 0.263, *P* < 0.05); and (2) disease versus healthy controls (|logLJFC| > 0.5, adj.*P* < 0.05). The resulting sets of markedly dysregulated genes were designated as differentially expressed genes DEGs1 and DEGs2, respectively. The candidate genes for downstream analysis were identified by determining the common genes shared among DEGs1, DEGs2, and AAMRGs. Gene Ontology (GO) and Kyoto Encyclopedia of Genes and Genomes (KEGG) pathway enrichment analyses were executed for the identified candidate genes via the ClusterProfiler (v4.6.2) (Yu et al. 2012) (adj.*P* < 0.05).

### 2.4 Machine learning

Candidate genes underwent feature selection through three distinct computational approaches implemented in R: (i) least absolute shrinkage and selection operator (LASSO) regression via glmnet (v4.1.10) (Friedman et al. 2010) with 10-fold cross-validation; (ii) support vector machine–based recursive feature elimination (SVM-RFE) applying caret (v6.0-94) (Shi et al. 2023), also employing 10-fold cross-validation; and (iii) the Boruta algorithm via Boruta (v8.0.0) (Maurya et al. 2023). Genes consistently identified across all three methods were designated as high-confidence candidate biomarkers. Their expression patterns—upregulation or downregulation—and distinguishing performance, which was measured by the area under the receiver operating characteristic (ROC) curve (AUC), were then assessed independently in two independent cohorts: GSE14323 and GSE84044. Biomarkers were ultimately defined as those exhibiting both AUC > 0.7 in both datasets and concordant directional expression changes (consistently up- or down-regulated) across the two cohorts (*P* < 0.05).

### 2.5 Immune infiltration analysis

The GSE14323 cohort’s immune cell infiltration levels were measured employing the CIBERSORT computational approach. The immune cell compositions of the cluster2 and cluster1 amino acid metabolism subtypes, as well as the LF and control groups, were compared. To evaluate any relationships between the detected biomarkers and immune cell types displaying notable infiltration variations between the disease and control groups, Spearman’s rank correlation coefficient was also computed.

### 2.6 Enrichment analysis

Initially, the Hallmark gene set was retrieved from the MSigDB. In the GSE14323 dataset, we computed Spearman correlation coefficients between each biomarker and all expressed genes via the psych (v2.4.3). Subsequently, gene set enrichment analysis (GSEA) was executed on the ranked correlation list by adopting clusterProfiler (|NES| > 1, adj.*P* < 0.05), as well as on the ranked DEGs1 list. Concurrently, pathway activity differences between amino acid metabolism–defined subtypes (cluster2 vs. cluster1) were assessed by adopting gene set variation analysis (GSVA) (v1.46.0) (Gui et al. 2024) integrated with the limma (v3.54.2), leveraging the identical Hallmark gene set (|t| > 2, adj.*P* < 0.05).

### 2.7 scRNA-seq analysis

scRNA-seq data preprocessing was implemented by adopting the Seurat (v4.4.0) (Hao et al. 2021). Quality control excluded cells based on the following criteria: nFeature_RNA ≤ 300 or ≥ 3,000, nCount_RNA ≥ 10,000; and mitochondrial gene expression proportion (percent.mt) ≥ 10%. Following log-normalization, highly variable genes (HVGs) were detected via VST, and the 2,000 genes exhibiting the greatest variability were selected for subsequent analyses. DoubletFinder (v2.0.4) (She et al. 2025) was applied to detect and remove potential doublets. Principal component analysis (PCA) was conducted, and the elbow plot of cumulative variance explained by each principal component was generated to guide dimension selection; the first 30 principal components (PCs) were retained for subsequent analyses. Unsupervised clustering was executed by applying Seurat’s FindNeighbors and FindClusters functions (resolution = 0.4), and cell embeddings were visualized in two-dimensional space via Uniform Manifold Approximation and Projection (UMAP). Cell type annotation was carried out by integrating canonical marker gene expression profiles with published literature (Huang et al. 2023; Luo et al. 2024; Ramachandran et al. 2019). Differential expression of candidate biomarkers across annotated cell types was assessed between disease and control conditions; cell types exhibiting both elevated baseline expression and statistically significant intergroup differences were designated as key cells. Intercellular communication was inferred by applying CellChat (v1.6.1) (Jin et al. 2021), focusing on ligand–receptor interaction strength and signaling specificity. Finally, pseudotime trajectory inference for key cell populations was implemented by adopting Monocle3 (v2.26.0) (Trapnell et al. 2014), and the dynamic expression patterns of biomarkers across developmental branches were visualized along the inferred trajectories.

### 2.8 Regulatory network

Potential miRNAs of biomarkers were identified in the starBase platform (https://rnasysu.com/encori/). Interactions between these miRNAs and their putative lncRNA targets (clipExpNum ≥ 18) were also extracted from starBase. Transcription factors (TFs) regulating the identified biomarker genes were obtained from the ChEA3 resource (https://maayanlab.cloud/chea3/). Subsequently, two regulatory networks were built: a multi-layered mRNA–miRNA–lncRNA interaction network, and a TF–mRNA network.

### 2.9 Drug prediction and molecular docking

Drug candidates targeting the identified biomarkers were prioritized in the DSigDB database (https://dsigdb.tanlab.org/; *P* < 0.01). Among the top-ranked compounds, those with favorable safety profiles and polypharmacological potential were selected for further investigation. Their three-dimensional structures (SDF format) were retrieved from PubChem (https://pubchem.ncbi.nlm.nih.gov/), while the high-confidence predicted protein structures of the biomarker targets were obtained from the Protein Data Bank (https://www.rcsb.org/; PDB format). Prior to docking, all PDB files were preprocessed in PyMOL (v2.5, https://pymol.org/) to remove crystallographic water molecules, co-crystallized ligands, and non-essential heteroatoms. Molecular docking simulations were performed using the gnina (v0.1.0.0) (McNutt et al. 2021). Docking poses were evaluated based on binding affinity (kcal/mol) and interaction topology, and representative complexes were visualized and annotated using PyMOL.

### 2.10 Hepatic stellate cell culture and fibrosis model establishment

The human hepatic stellate cell line LX-2, a well-established immortalized human hepatic stellate cell line for fibrosis research, was purchased from Vetcell Biotechnology (China, Cat# VCH00014) (Xu et al. 2005). Cells were cultured in DMEM medium supplemented with 10% fetal bovine serum (FBS) and 1% penicillin-streptomycin (P/S) at 37°C in a humidified atmosphere containing 5% CO2. Before stimulation, cells were serum-starved in DMEM containing 1% FBS for 24 h. For fibrosis model establishment and subsequent validation, cells were divided into two groups: the control group received no treatment, whereas the model group was stimulated with TGF-β1 (10 ng/mL; MCE, HY-P70543/10ug) for 24 h to induce LX-2 activation, as TGF-β signaling is a central driver of hepatic stellate cell activation and TGF-β1-treated LX-2 cells exhibit transcriptional changes consistent with fibrogenic activation (Dewidar et al. 2019; Carson et al. 2021).

### 2.11 Cell viability assay

After trypsinization and counting, LX-2 cells in the logarithmic growth phase were seeded onto 96-well plates (3-4×103 cells/well). After cell attachment, cells were treated with varying doses of TGF-β1 (0, 1, 5, 10, 20, 40, 80, and 100 ng/mL) for 24 h. Subsequently, 10 μL CCK-8 solution was added to each well and incubated for 1 h. A microplate reader was used to measure absorbance at 450 nm, and 10 ng/mL TGF-β1 was selected for subsequent experiments based on the cell viability results.

### 2.12 Reverse transcription–quantitative PCR (RT-qPCR)

The FastPure Complex Tissue/Cell Total RNA Isolation Kit (Vazyme Biotech, Cat. RC113-01) was employed to isolate total RNA from cultivated cells. Reverse transcription was performed with ABScript III RT Master Mix with integrated gDNA removal (ABclonal) on high-quality RNA, which was defined by an OD260/OD280 ratio > 1.8 as determined by NanoDrop 500 spectrophotometry. Quantitative PCR amplification was then carried out with the Genious 2× SYBR Green Fast RT-qPCR Mix (ABclonal), and each sample was analyzed in triplicate. GAPDH served as the endogenous control. The expression levels of GSTP1, LDHB, OXCT1, PTGDS, ACTA2, and COL1A1 were quantified, with primer sequences listed in Table S1. ACTA2 and COL1A1 were included as classical fibrogenic markers to verify successful activation of LX-2 cells after TGF-β1 stimulation (Dewidar et al. 2019; Carson et al. 2021). Relative gene expression levels were calculated using the 2^–ΔΔCt method.

### 2.13 Statistical analysis

All bioinformatic analyses were performed using R software. Experimental data are presented as mean ± SD. Comparisons between two groups were performed using an unpaired Student’s t-test, whereas multiple-group comparisons were conducted using one-way analysis of variance. A two-sided P < 0.05 was considered statistically significant.

## 3 Results

### 3.1 Identification of 67 high-priority candidate genes in LF

Based on amino acid metabolism-related gene (AAMRGs) expression, the 41 LF samples in GSE14323 were stratified into two distinct subtypes, Cluster 1 and Cluster 2 (**Fig. 1A**). Subsequent analysis revealed divergent immune microenvironments and functional pathway activities between the two groups. Specifically, 11 immune cell types exhibited significantly higher infiltration levels in Cluster 2, including activated CD8^+^ T cells and natural killer T cells (**Fig. 1B**). Gene set variation analysis (GSVA) based on the HALLMARK gene set identified 15 differential enriched pathways (**Fig. 1C**). Notably, Cluster 2 displayed marked activation of the Wnt/β-catenin signaling and apical junction pathways, whereas Cluster 1 was enriched in xenobiotic metabolism and adipogenesis. Gene set enrichment analysis (GSEA) corroborated these findings, identifying 25 significantly enriched pathways (**Fig. 1D, Table S2**). Cluster 2-associated pathways included allograft rejection and apical junction, while Cluster 1 maintained a strong association with metabolic processes such as xenobiotic and fatty acid metabolism. These results indicate that the Cluster 2 exhibits stronger immune activation and distinct signaling pathway engagement, whereas Cluster 1 was more strongly associated with metabolic pathways. A total of 2,057 DEGs1 were identified between the two subtypes, comprising 780 upregulated and 1,277 downregulated genes (**Fig. 1E-F**). In the GSE14323 dataset, comparison between the disease cohort and the healthy controls yielded 2,157 DEGs2, of which 1,334 were upregulated and 823 downregulated (**Fig. 1G-H**). By intersecting DEGs1, DEGs2 and AAMRGs, we refined the focus to a set of 67 high-priority candidate genes (**Fig. 1I**). Functional annotation via GO of these candidates revealed marked enrichment across 303 terms, predominantly linked to carboxylic acid biosynthesis, aromatase activity, and mitochondrial matrix localization (**Fig. 1J, Table S3**). Pathway-level analysis using KEGG further identified 51 statistically enriched pathways, notably including arginine biosynthesis and the metabolism of alanine, aspartate, and glutamate—key processes within amino acid metabolic networks (**Fig. 1K, Table S3**).

**Fig. 1.**
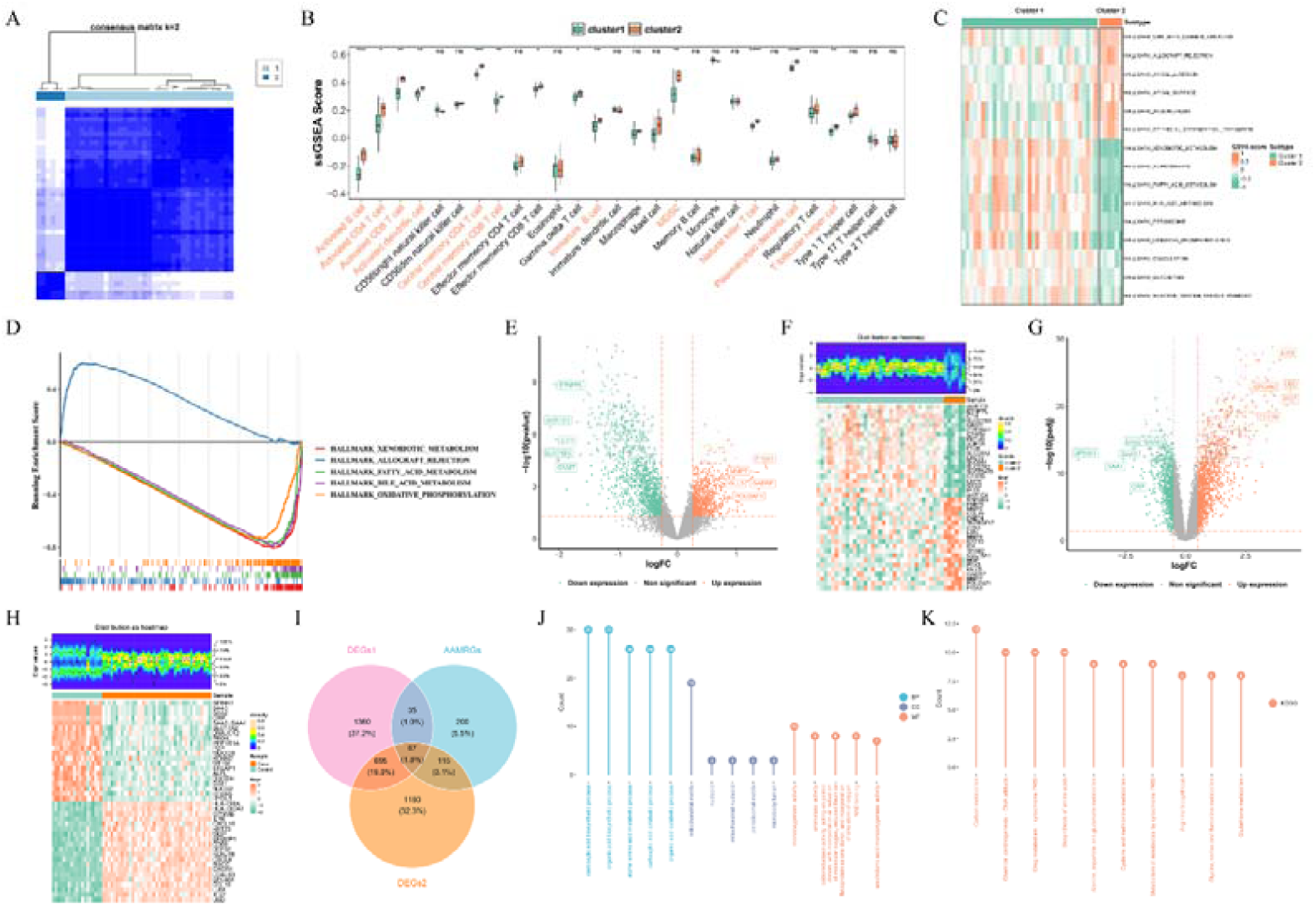
Identification of candidate genes. (**A**) Consensus clustering of liver fibrosis patients in GSE14323 dataset based on amino acid metabolism-related genes (AAMRGs) expression. (**B**) The difference in the proportion of immune cells between two subtypes. (**C**) Gene set variation analysis (GSVA) of two subtypes. (**D**) Gene set enrichment analysis (GSEA) of differentially expressed genes (DEGs)1 between two subtypes. (**E**) Volcano plot of DEGs1 between two subtypes. (**F**) Heatmap of top 20 up-regulated and all down-regulated DEGs1. (**G**) Volcano plot of DEGs2 between liver fibrosis samples and control samples in GSE14323 dataset. (**H**) Heatmap of top 20 up/down-regulated DEGs2. (**I**) Venn diagram of two sets of DEGs with AAMRGs. (**J**) Top 5 terms in Gene Ontology (GO) functions and Kyoto Encyclopedia of Genes and Genomes (KEGG) enriched by 67 candidate genes. (**K**) Top 10 Kyoto Encyclopedia of Genes and Genomes (KEGG) pathways enriched by 67 candidate genes. ns: not significant, * *P* < 0.05, ** *P* < 0.01, *** *P* < 0.001, **** *P* < 0.0001

### 3.2 Identification of GSTP1, LDHB, OXCT1, and PTGDS as biomarkers

To further refine candidate selection, three distinct feature-selection algorithms were applied. LASSO regression (with λ.min = 0.005) identified 14 feature genes, while SVM-RFE and Boruta selected 54 and 40 feature genes, respectively (**Fig. 2A-C**). Intersection analysis across these three methods revealed a core set of 13 overlapping genes: PTGDS, LDHB, UGDH, GSTP1, IDH2, GLUD1, NADK, OXCT1, MAOA, CYP1B1, HADHB, GOT1, and HPD (**Fig. 2D**). Subsequent expression profiling demonstrated that among these candidates, GSTP1, LDHB, OXCT1, and PTGDS were consistently upregulated in LF tissues (**Fig. 2E-F**). Moreover, these four genes displayed robust diagnostic performance, achieving AUC values exceeding 0.7 in both GSE14323 and GSE84044 cohorts. Based on these findings, they were selected as the final diagnostic biomarker panel for LF (**Fig. 2G-H**).

**Fig. 2.**
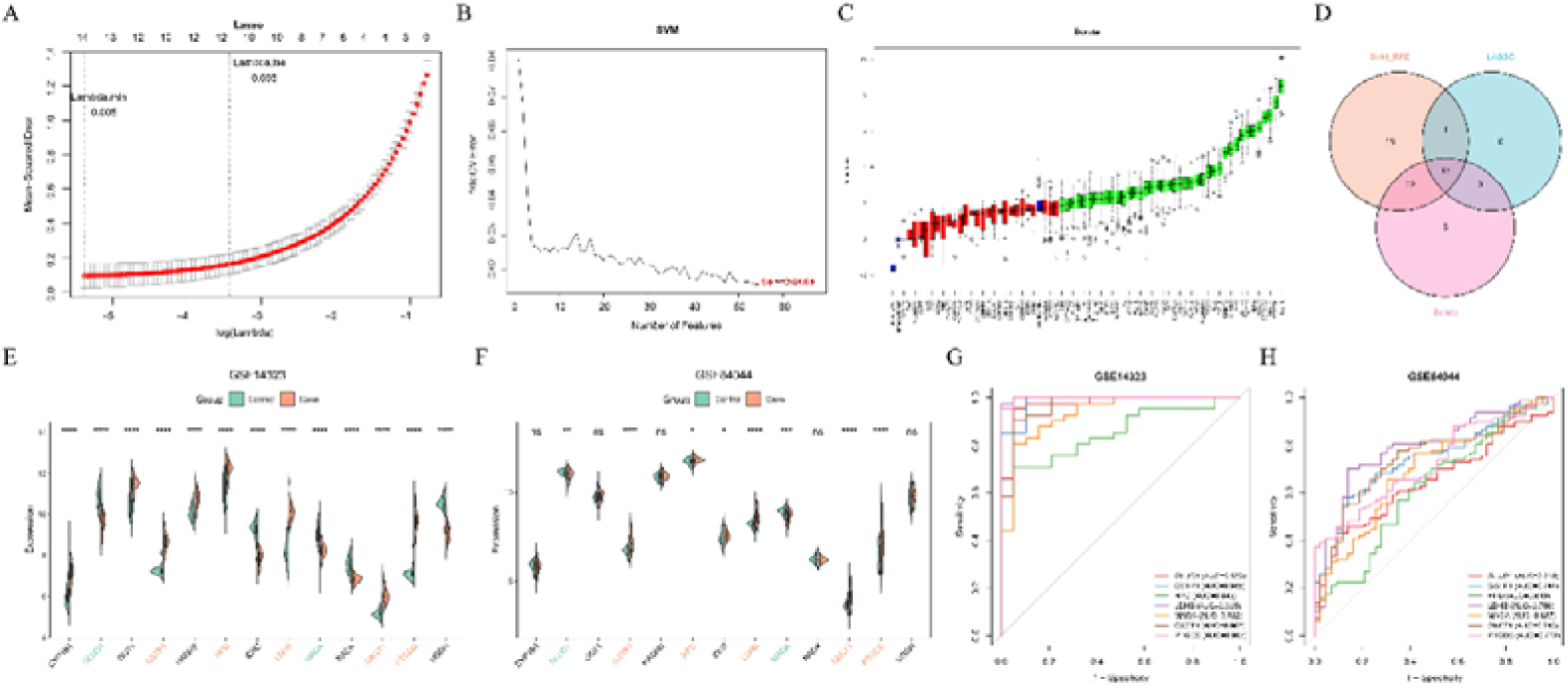
Identification of GSTP1, LDHB, OXCT1, and PTGDS as biomarkers. (**A**) Least absolute shrinkage and selection operator (LASSO) regression in the GSE14323 cohort. (**B**) Support vector machine–based recursive feature elimination (SVM-RFE) in the GSE14323 cohort. (**C**) Boruta in the GSE14323 cohort. (**D**) Venn diagram of feature genes in three machine learning models. (**E**) Candidate biomarker expression in the GSE14323 cohort. (**F**) Candidate biomarker expression in the GSE84044 cohort. (**G**) Receiver operating characteristic (ROC) curve in the GSE14323 cohort of candidate biomarkers with concordant directional expression changes. (**H**) Receiver operating characteristic (ROC) curve in the GSE84044 cohort of candidate biomarkers with concordant directional expression changes. ns: not significant, * *P* < 0.05, ** *P* < 0.01, *** *P* < 0.001, **** *P* < 0.0001

### 3.3 Immune-related functional signatures and cellular correlates of four fibrosis-associated biomarkers

Functional enrichment analysis identified 28-31 biological pathways associated with each of the four identified biomarkers (GSTP1, LDHB, OXCT1, and PTGDS), with detailed pathway annotations provided in Supplementary Table S4. They were collectively involved in epithelial–mesenchymal transition (EMT), interferon signaling, and key immune-regulatory pathways, including TNFα/NF-κB (**Fig. 3A-D, Table S4**). To assess their immunological relevance, comprehensive immune infiltration profiling was performed. This analysis revealed statistically significant differences in the relative abundance of 12 distinct immune cell subsets between LF patients and healthy controls (**Fig. 3E-F**). Specifically, M1 macrophages, γδ T cells, and resting memory CD4LJ T cells were markedly enriched in the fibrotic cohort and showed robust positive correlations with all four biomarkers. In contrast, follicular helper T cells (Tfh) and regulatory T cells (Tregs) displayed inverse relationships with these biomarkers. Correlation quantification further demonstrated that LDHB exhibited the strongest positive correlation with CD4 memory quiescent T cells (r = 0.73) and the most pronounced negative correlation with Tregs (r = −0.85) (**Fig. 3G**). Collectively, these findings suggested that the identified biomarkers may modulate the hepatic immune landscape during fibrosis progression by influencing the recruitment and activation of specific immune subsets.

**Fig. 3.**
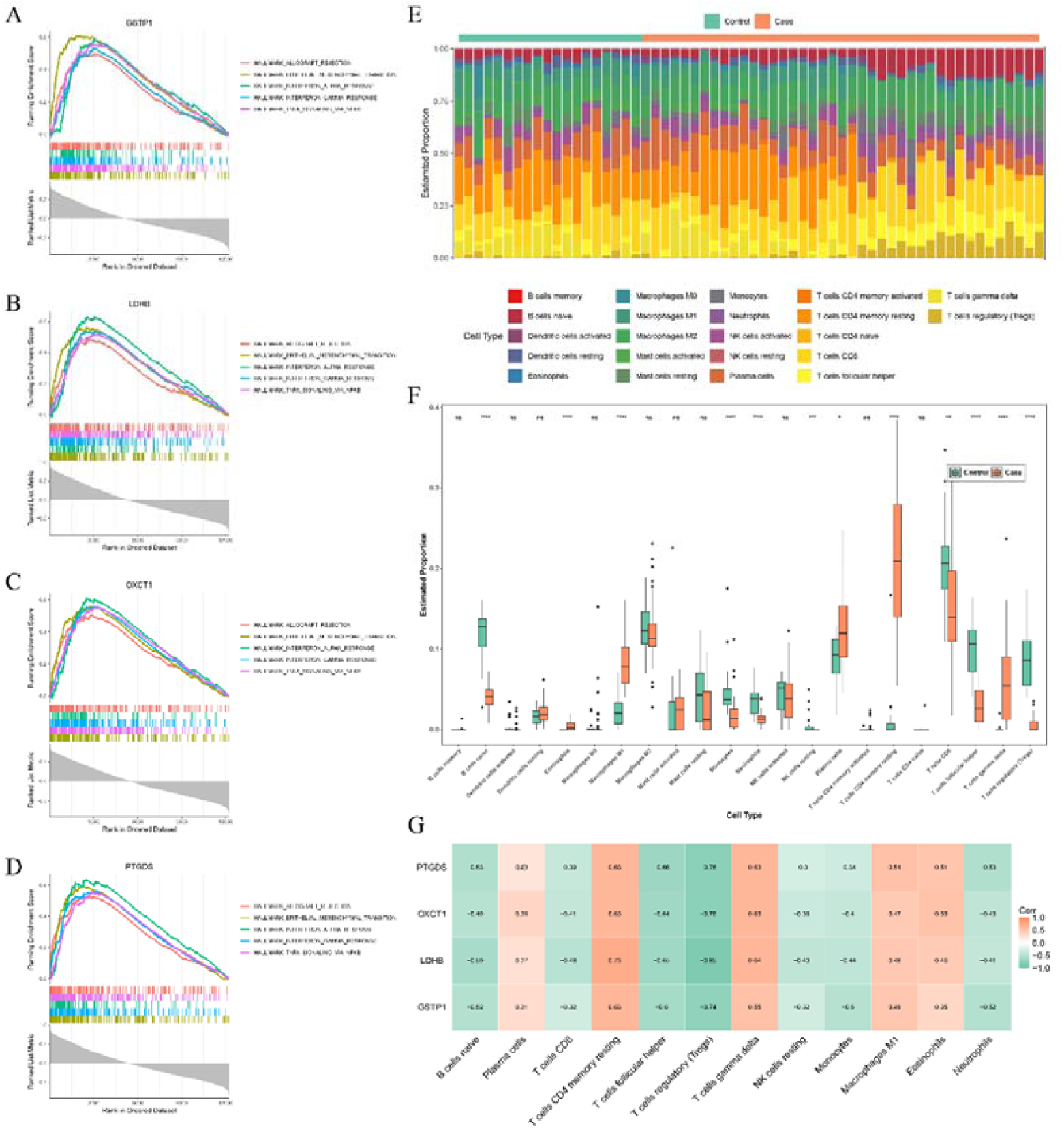
GSEA and the correlation with immune cells of biomarkers. (**A**) GSEA of GSTP1. (**B**) GSEA of LDHB. (**C**) GSEA of OXCT1. (**D**) GSEA of PTGDS. (**E**) Stacked chart of the proportion of immune cell infiltration in GSE14323 dataset. (**F**) The difference in the proportion of immune cells between liver fibrosis samples and control samples in GSE14323 dataset. (**G**) The correlation between immune cells and biomarkers. ns: not significant, * *P* < 0.05, ** *P* < 0.01, *** *P* < 0.001, **** *P* < 0.0001

### 3.4 Single-cell landscape of biomarker expression and cholangiocyte-centric interactions in LF

Subsequently, the specific expression patterns of the four biomarkers at the single-cell level were examined. After quality control, double-cell removal, dimension reduction, and unsupervised clustering of the GSE136103 dataset, 51,298 cells were partitioned into 22 cell clusters (**Fig. 4A, Fig. S1A-E**). A total of 10 types of cells were annotated, namely: T cells, macrophages, endothelial cells, B cells, cholangiocytes, mesenchymal cells, cycling cells, dendritic cells, plasma cells, and mast cells (**Fig. 4B, Fig. S1F**). Cell-type-specific expression profiling revealed distinct distribution patterns of the biomarkers. GSTP1 exhibited elevated expression levels in cholangiocytes, cycling cells, and macrophages. LDHB showed pronounced expression in mast cells, dendritic cells, cholangiocytes, and mesenchymal cells. OXCT1 was predominantly expressed in cycling cells, whereas PTGDS displayed strong enrichment specifically in dendritic cells. Notably, both OXCT1 and PTGDS were also detected in cholangiocytes (**Fig. 4C**). Importantly, cholangiocytes consistently expressed all four biomarkers, and the collective expression levels of these biomarkers differed significantly among the experimental groups (**Fig. 4D**). Based on these findings, cholangiocytes were selected as the key cell type for downstream investigations. To investigate the cellular context of these biomarkers, we performed a comprehensive analysis of intercellular communication networks concentrated on cholangiocytes. Ligand–receptor interaction mapping visualized robust signaling crosstalk between cholangiocytes and both mesenchymal cells and macrophages, primarily mediated by the MIF–CD74/CXCR4 axis and the APP–CD74 interaction (**Fig. 4E-F**). Pseudotime trajectory of cholangiocyte development identified five distinct differentiation states. In the disease cohort, cells were predominantly found in states 3 through 5 (**Fig. 4G**). Biomarker expression profiling along this Pseudotime demonstrated that four markers were consistently detectable across the trajectory. Notably, GSTP1 and LDHB exhibited significantly elevated expression levels specifically in states 3–5 (**Fig. 4H**).

**Fig. 4.**
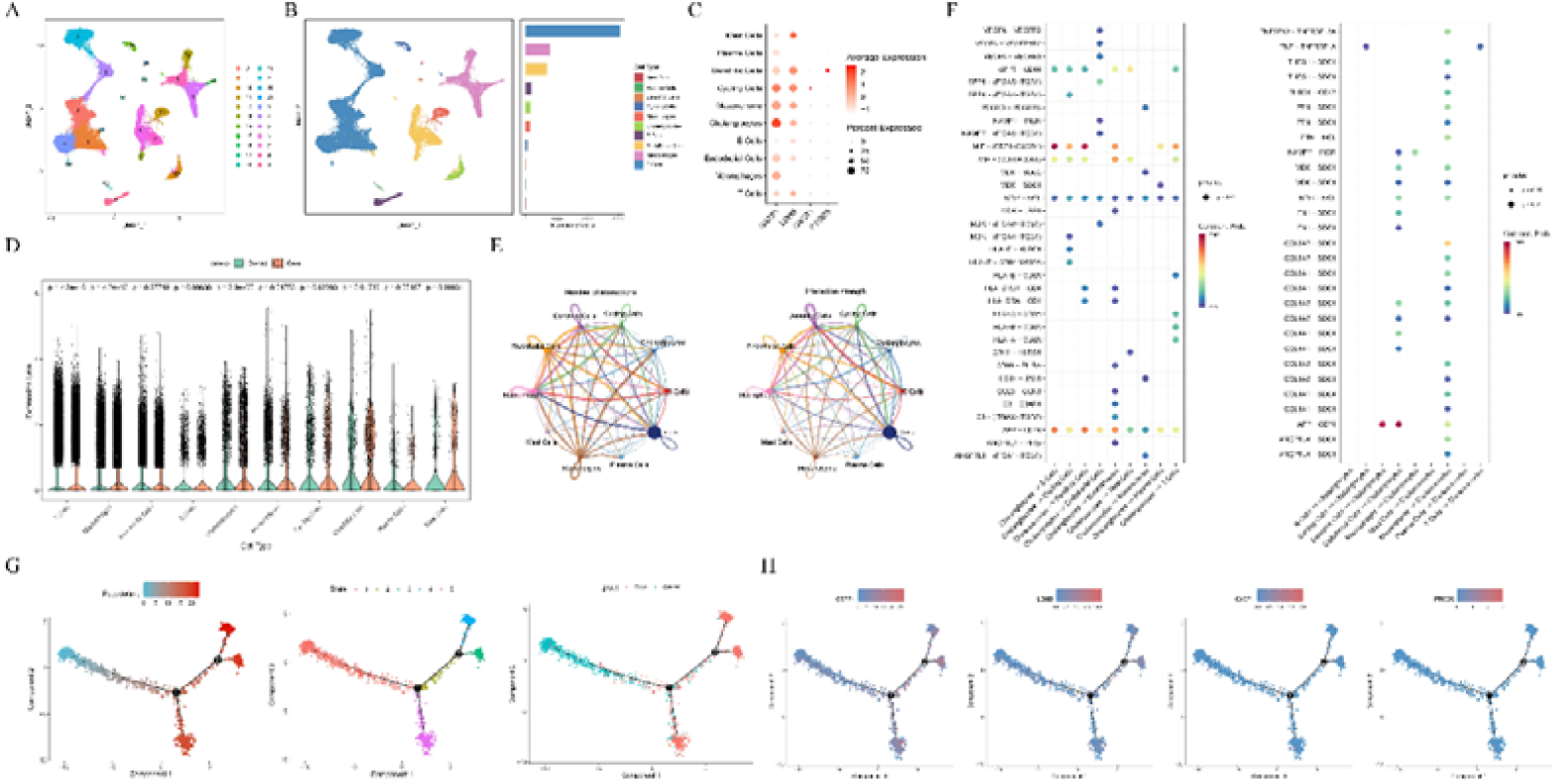
Identification of key cellular type in liver fibrosis. (**A**) Uniform Manifold Approximation and Projection (UMAP) clustering plot of 22 clusters in the single-cell RNA sequencing (scRNA-seq) dataset. (**B**) UMAP plot illustrating the distribution of 10 cell types and the cellular composition of each cell type. (**C**) The expression of biomarkers in each cell type. (**D**) The difference in overall biomarker expression between liver fibrosis and control groups. (**E**) Cellular communication network illustrating the number (left) and strength (right) of interactions among cell types. (**F**) Signaling patterns of cell type interactions. (**G**) Pseudotime trajectory analysis; from left to right: Time trajectory of cholangiocytes; Cell state distribution of cholangiocytes in pseudotime trajectory analysis; Time trajectory of cholangiocytes in the liver fibrosis group and the control group. (**H**) The expression of biomarkers in time trajectory of cholangiocytes

### 3.5 Upstream regulatory molecules and targeted drugs of biomarkers

To elucidate the potential regulatory mechanisms governing the four biomarkers (GSTP1, LDHB, OXCT1, and PTGDS), we established both mRNA–miRNA–lncRNA and TF–mRNA regulatory networks. The mRNA–miRNA–lncRNA network comprised 4 mRNAs, 98 miRNAs, and 19 lncRNAs. Notably, this analysis suggested that lncRNAs such as AC010980.2, MALAT1, and NORAD may modulate OXCT1 expression through competitive binding to hsa-miR-3163 (**Fig. 5A**). Concurrently, a TF–mRNA regulatory network comprising 4 mRNAs and 72 TFs was established (**Fig. 5B**). Among the biomarkers, GSTP1 was predicted to be targeted by 22 TFs, LDHB by 20 TFs, OXCT1 by 30 TFs, and PTGDS by 14 TFs. Intriguingly, several TFs were found to co-regulate multiple biomarkers: NANOG, E2F1, and E2F4 were shared by LDHB and GSTP1; RELA co-regulated GSTP1 and PTGDS; RUNX1 targeted both LDHB and PTGDS; WT1, AR, and SMAD4 jointly regulated OXCT1 and LDHB; and TET1, SOX9, MEIS1, MYC, and MITF were common to OXCT1 and PTGDS. Drug prediction was implemented to identify existing pharmaceutical agents that could interact with the identified biomarkers. The screening identified 86 candidate drugs associated with GSTP1, 3 with LDHB, 2 with OXCT1, and 4 with PTGDS (**Fig. 5C**). Notably, 5-fluorouracil emerged as a multi-target compound and was predicted by DSigDB to interact with three biomarkers, namely GSTP1, LDHB, and OXCT1. It was therefore selected for subsequent molecular docking analysis against all four identified biomarkers. The calculated binding affinities of 5-fluorouracil with GSTP1, LDHB, OXCT1, and PTGDS were −5.13, −5.07, −6.34, and −4.97 kcal/mol, respectively. Among them, OXCT1 showed the strongest predicted binding affinity, and the 5-fluorouracil–OXCT1 complex was selected as the representative docking model in **Fig. 5D**.

**Fig. 5.**
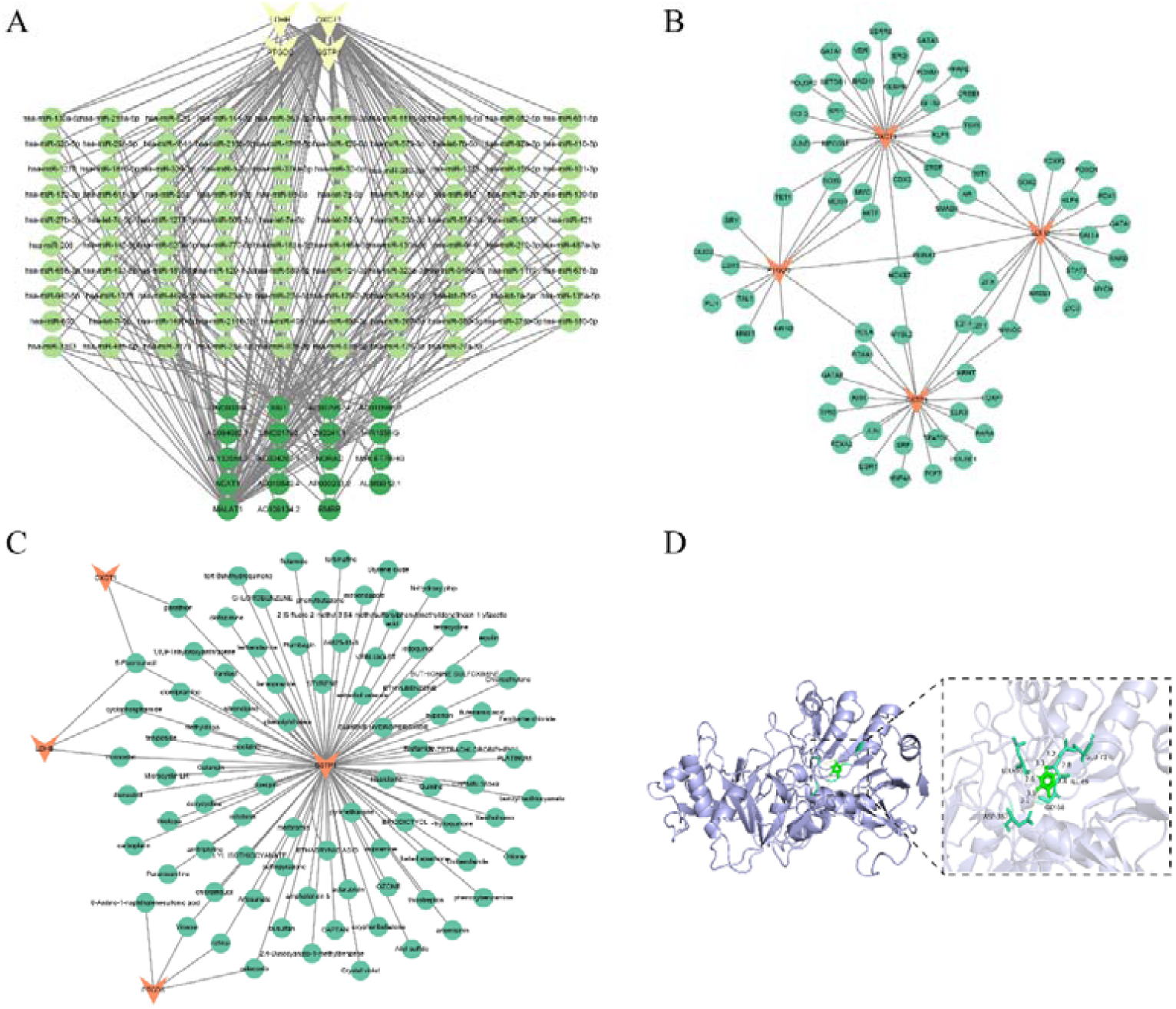
Regulatory network and potential therapeutic agents of biomarkers. (**A**) mRNA-miRNA-lncRNA regulatory network. (**B**) TF–mRNA regulatory network. (**C**) Gene-drug network. (**D**) Molecular docking of 5-fluorouracil and OXCT1

### 3.6 Validation of biomarker expression in the TGF-**β**1-induced LX-2 fibrosis model

To experimentally validate the expression patterns of the four identified biomarkers (GSTP1, LDHB, OXCT1, and PTGDS) in LF, we established an in vitro fibrosis model using the human hepatic stellate cell line LX-2. A concentration-response experiment was first performed to select an appropriate TGF-β1 dose. The CCK-8 assay showed that treatment with 10 ng/mL TGF-β1 yielded the highest cell viability and was therefore selected for subsequent modeling (Fig. 6A). To verify successful model establishment, the expression of the classical fibrogenic markers ACTA2 and COL1A1 was further examined by RT-qPCR. Both ACTA2 and COL1A1 were significantly upregulated in the TGF-β1-treated group, confirming successful activation of LX-2 cells and establishment of the in vitro fibrotic model (Fig. 6F–G). On this basis, the expression levels of the four candidate biomarkers were further assessed. Consistent with the bioinformatics analysis, GSTP1 and LDHB were notably upregulated in the TGF-β1-treated LX-2 cells compared with controls. However, OXCT1 and PTGDS displayed opposite expression trends and were decreased following TGF-β1 stimulation (Fig. 6B-E).

**Fig. 6.**
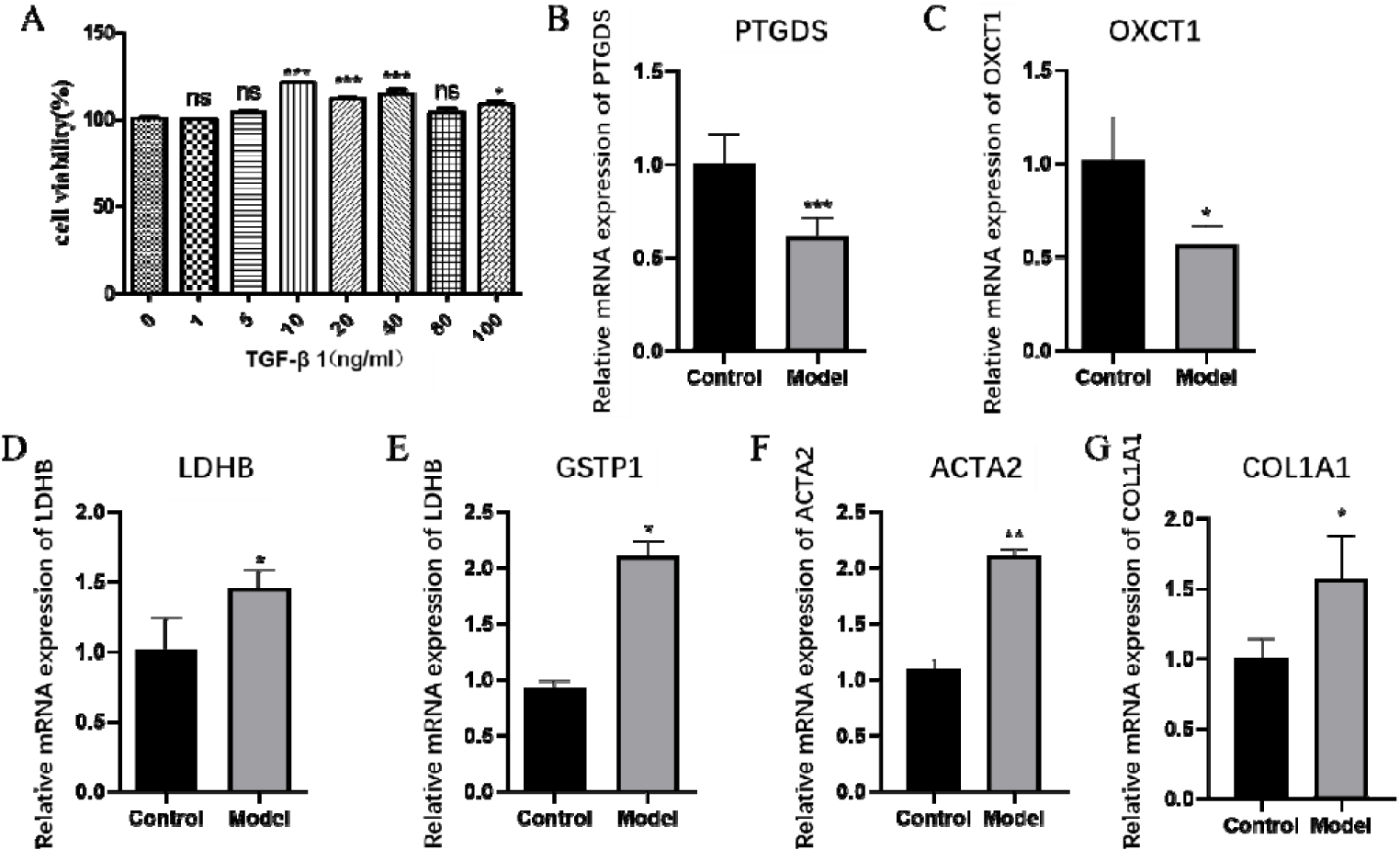
Validation of biomarker expression in the TGF-β1-induced LX-2 fibrosis model. (**A**) Cell viability of LX-2 cells treated with different concentrations of TGF-β1 (0–100 ng/mL). (**B**) RT-qPCR analysis of PTGDS in control and model cells. (**C**) RT-qPCR analysis of OXCT1 in control and model cells. (**D**) RT-qPCR analysis of LDHB in control and model cells. (**E**) RT-qPCR analysis of GSTP1 in control and model cells. (**F**) RT-qPCR analysis of ACTA2 in control and model cells. (**G**) RT-qPCR analysis of COL1A1 in control and model cells. ns: not significant, * P < 0.05, ** P < 0.01, *** P < 0.001.

## 4 Discussion

LF is increasingly understood as a dynamic multicellular process characterized not only by extracellular matrix accumulation, but also by profound metabolic rewiring, inflammatory amplification, and niche-dependent intercellular communication. Recent work has highlighted that metabolic adaptation is not a secondary epiphenomenon, but a core driver of fibrogenic activation in hepatic stellate cells, macrophages, and other non-parenchymal populations (Horn and Tacke 2024; Gilgenkrantz et al. 2021). In this study, we identified molecular subtypes associated with amino acid metabolism in liver fibrosis and screened a set of core biomarkers—including GSTP1, LDHB, OXCT1, and PTGDS—that are closely linked to immune and remodeling-related pathways. Single-cell analysis further revealed that these genes are highly enriched in cholangiocytes, suggesting that this cell type may play a previously underappreciated key role in the pathogenesis of liver fibrosis. Importantly, the TGF-β1-induced LX-2 model used for wet-lab validation was supported by significant upregulation of ACTA2 and COL1A1, indicating successful fibrotic activation at the mRNA level. Within this validated model, GSTP1 and LDHB were directionally consistent with the bioinformatics results. Taken together, these findings suggest that amino acid metabolism-related transcriptional remodeling is closely coupled to immune heterogeneity and cell-type-specific communication in LF, and may provide a biologically informative basis for molecular stratification.

A major finding of this study is that LF samples could be divided into two AAM-related molecular clusters with distinct biological characteristics. Compared with cluster1, cluster2 showed stronger enrichment of immune activation, apical junction, and Wnt/β-catenin signaling, whereas cluster1 retained relatively higher xenobiotic and fatty acid metabolic features. This pattern is biologically meaningful because aberrant Wnt/β-catenin activation has been repeatedly implicated in chronic liver injury, hepatic stellate cell activation, hepatobiliary remodeling, and progression from steatohepatitis to fibrosis (Miao et al. 2013; Shree Harini and Ezhilarasan 2023). Combining the above AAM clustering with the Wnt/β-catenin signaling pathway, our results suggest that molecular typing based on amino acid metabolism may capture fibrotic endotypes with biological significance, rather than random transcriptome variations. From a translational perspective, such clustering may be useful because a metabolically biased subtype and an immune-remodeling-dominant subtype are unlikely to respond identically to future antifibrotic or immunometabolic interventions. Although our datasets did not allow direct correlation with fibrosis stage or etiology, the present analysis still suggests that AAM-related stratification may refine disease heterogeneity in a way that conventional histology alone cannot fully resolve.

Among the four genes, GSTP1 and LDHB both were consistently upregulated in fibrotic tissues, retained diagnostic value across datasets, and were directionally supported by the LX-2 validation model. GSTP1 is a glutathione S-transferase with a well-established role in detoxification and oxidative stress adaptation; recent liver work further showed that loss of GSTP1 stability promotes reactive oxygen species accumulation, inflammatory cytokine production, and liver injury, supporting its close connection to oxidative-stress-sensitive hepatic homeostasis (Zhong et al. 2024). In the context of LF, where oxidative stress and redox imbalance are central permissive factors for stellate-cell activation and inflammatory amplification, GSTP1 upregulation may reflect a compensatory but insufficient stress-adaptive response. LDHB, in contrast, links the AAM signature to lactate-centered metabolic plasticity. LDHB catalyzes the reversible conversion between lactate and pyruvate and thereby participates in determining whether lactate is accumulated or reutilized; lactate metabolism has increasingly been recognized as relevant to LF and broader immune dysfunction (Yao et al. 2024). Our results showed that LDHB had the strongest inverse correlation with Tregs and a positive association with M1 macrophage-related immune features, suggesting that it may mark a microenvironment in which altered lactate handling coexists with impaired immunoregulatory restraint. Although the immunosuppressive consequences of lactate have been well characterized in tumor biology, accumulating evidence in liver research indicates that abnormal lactate metabolism also contributes to fibrogenesis and inflammatory remodeling (Brand et al. 2016; Yao et al. 2024). These findings position GSTP1 and LDHB as functionally interconnected biomarkers that bridge oxidative stress adaptation and lactate-centered metabolic reprogramming, two complementary drivers of the fibrotic microenvironment that may jointly sustain hepatic stellate cell activation and immune dysregulation in LF progression.

The biological interpretation of OXCT1 and PTGDS requires additional nuance. OXCT1 is the rate-limiting enzyme for ketone body utilization and therefore reflects the capacity of cells to engage in ketolysis and alternative oxidative substrate use. Although direct evidence linking OXCT1 to LF remains limited, emerging studies suggest that OXCT1-dependent ketone metabolism can shape the functional states of specific immune populations, including tumor-associated macrophages (TAMs) and CD8+ T cells (Zhu et al. 2024), supporting its relevance as an immunometabolic regulator rather than a purely housekeeping enzyme. In the context of our data, OXCT1 may therefore represent a marker of altered substrate preference and metabolic adaptability in selected fibrotic cell states rather than a universal marker of stellate-cell activation. PTGDS, which catalyzes prostaglandin D2 synthesis, provides a different layer of interpretation. Rather than directly indexing canonical fibrogenic activation, PTGDS more plausibly links the biomarker panel to lipid mediator remodeling and immune-cell-associated signaling (Shimura et al. 2010). This interpretation is consistent with our single-cell analysis, in which PTGDS showed prominent enrichment in dendritic cells and was also detectable in cholangiocytes, suggesting that it may participate in local paracrine inflammatory-immune regulation within the fibrotic niche rather than representing a universal intracellular fibrosis program. Importantly, the distinct biological roles of OXCT1 and PTGDS may also help explain why their expression trends in the LX-2 model were not fully concordant with the transcriptomic findings. In this sense, the four-gene signature should be understood as a composite network marker in which the individual genes contribute non-identical but complementary information. This also explains why a multigene panel is likely to outperform any single marker alone: LF is a heterogeneous process involving metabolic stress, inflammatory signaling, and cell-state diversity, and a combined signature is better positioned to improve diagnostic specificity and molecular stratification accuracy than a single-gene readout.

The single-cell results add an especially important mechanistic dimension by identifying cholangiocytes as the major population co-expressing all four biomarkers. This finding is biologically plausible and clinically relevant—cholangiocytes are no longer viewed as passive epithelial bystanders; rather, through ductular reaction, they participate in inflammatory recruitment, portal remodeling, and crosstalk with mesenchymal and immune populations in multiple chronic liver diseases (Mavila et al. 2024; Sato et al. 2023). In human cirrhosis, single-cell analysis has defined a specialized fibrotic niche composed of scar-associated macrophages, endothelial cells, mesenchymal cells, and epithelial populations, underscoring that fibrosis progression is sustained by niche-level cooperation rather than a single-cell program (Ramachandran et al. 2019). In our study, cholangiocytes not only concentrated the expression of all four AAM-related biomarkers, but also displayed active communication with macrophages and mesenchymal cells. Taken together, these findings support a cholangiocyte-centered communication hypothesis, in which amino acid metabolism-related remodeling is embedded within ductular reaction-associated fibrotic crosstalk. This interpretation is also consistent with recent literature indicating that ductular reaction is closely associated with hepatic inflammation and progressive fibrosis, and may serve as both a marker and mediator of portal fibrogenic remodeling (Mavila et al. 2024; Sato et al. 2023).

At the same time, the cholangiocyte-centered interpretation should be expressed with appropriate caution. Our data support cholangiocytes as a prominent co-expression compartment and a plausible signaling hub, but they do not prove that cholangiocytes are the sole initiating driver of fibrosis. This distinction is particularly important because the wet-lab validation was performed in TGF-β1-induced LX-2 cells, which model activated hepatic stellate cells rather than cholangiocytes. Although successful activation of the LX-2 model was supported by increased ACTA2 and COL1A1 expression, the cellular mismatch between the single-cell localization and the in vitro system likely explains why GSTP1 and LDHB were directionally validated whereas OXCT1 and PTGDS were not. Thus, the current validation should be interpreted as partial support for the biomarker panel rather than definitive confirmation of all four genes in the same cellular context. Future studies should prioritize cholangiocyte models, multicellular co-culture systems, organoids, or spatially resolved validation strategies to determine whether these genes function primarily within ductular cells, across fibrotic niches, or in cell-type-specific paracrine loops. This is particularly relevant given that current in vitro and ex vivo LF models each capture only part of the multicellular and etiologic complexity of human disease (Ros-Tarraga et al. 2025).

Compared with conventional fibrosis transcriptome studies that focus primarily on differential expression or generic pathway enrichment, our analysis specifically approached LF from the perspective of amino acid metabolism-related remodeling. This is meaningful because recent work increasingly places immunometabolic adaptation at the center of hepatic fibrogenesis (Horn and Tacke 2024; Gilgenkrantz et al. 2021). However, several limitations should nevertheless be acknowledged. First, the analyses were based mainly on retrospective public datasets, which inevitably introduce heterogeneity related to platform differences, sample processing, and incomplete clinical annotation. Second, the inferred immune infiltration patterns, regulatory networks, and molecular docking remain computational predictions and require direct functional validation. Third, the in vitro validation relied on a single stellate-cell model and thus could not fully represent the multicellular architecture of fibrotic liver tissue. Fourth, although the four-gene panel showed promising diagnostic performance, its value for fibrosis staging, etiologic subclassification, and therapeutic response prediction remains unknown. Despite these limitations, our study provides convergent evidence that amino acid metabolism-related signatures are closely linked to LF heterogeneity and may help bridge molecular diagnosis with mechanism-oriented stratification.

## 5 Conclusion

Overall, GSTP1, LDHB, OXCT1, and PTGDS constitute a biologically informative AAM-related biomarker set, and cholangiocyte-associated communication emerges as a plausible axis underlying fibrotic progression. These findings offer a rationale for future mechanistic and translational studies aimed at biomarker validation and metabolism-informed precision strategies in LF.

## Data Availability

Publicly available transcriptomic datasets analyzed in this study are available from the Gene Expression Omnibus (GEO) under accession numbers GSE14323, GSE84044, and GSE136103. Additional data generated in the present study are available from the corresponding author upon reasonable request.

https://www.ncbi.nlm.nih.gov/geo/query/acc.cgi?acc=GSE14323

https://www.ncbi.nlm.nih.gov/geo/query/acc.cgi?acc=GSE84044

https://www.ncbi.nlm.nih.gov/geo/query/acc.cgi?acc=GSE136103

