## Supplementary material for "Identification of amino acid metabolism-related biomarkers in liver fibrosis: a transcriptomic analysis with experimental validation": Supplementary Figure legend.docx

**Supplementary Information**

**Fig. S1** Single-cell RNA sequencing (scRNA-seq) data processing. (**A**) scRNA-seq data before (upper) and after (lower) quality control. (**B**) Selection of top 2,000 highly variable genes (HVGs). (**C**) Uniform Manifold Approximation and Projection (UMAP) plot illustrating the distribution of doublets in cells. (**D**) Permutation test. (**E**) Elbow plot of PCs. (**F**) Expression of marker genes in the main cell types
