## Supplementary figures and images for "Identification of amino acid metabolism-related biomarkers in liver fibrosis: a transcriptomic analysis with experimental validation"

### Fig. S1.tif

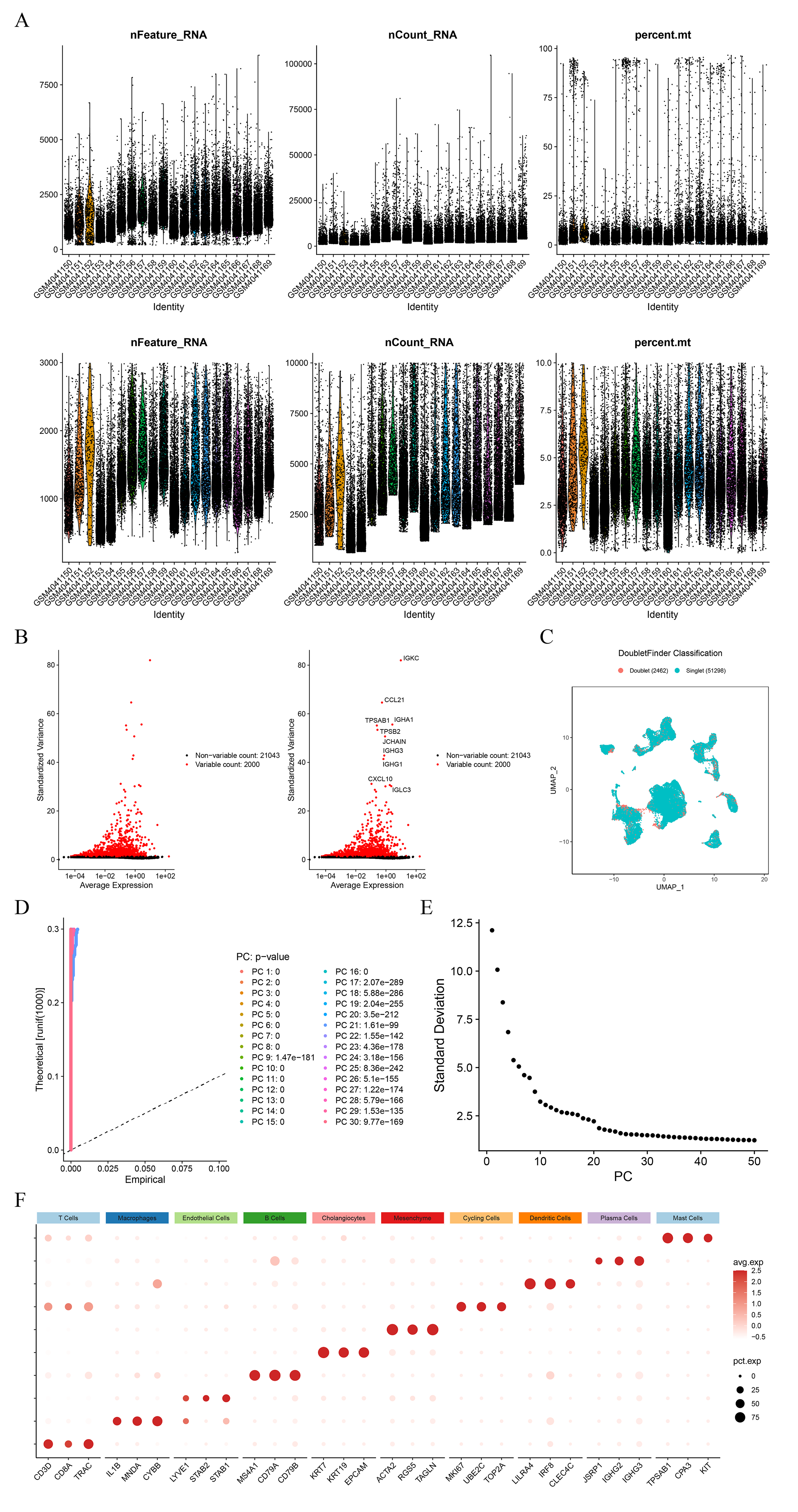
